# The relationship of COVID-19 severity with cardiovascular disease and its traditional risk factors: A systematic review and meta-analysis

**DOI:** 10.1101/2020.04.05.20054155

**Authors:** Kunihiro Matsushita, Ning Ding, Minghao Kou, Xiao Hu, Mengkun Chen, Yumin Gao, Yasuyuki Honda, David Dowdy, Yejin Mok, Junichi Ishigami, Lawrence J. Appel

## Abstract

**Background:** Whether cardiovascular disease (CVD) and its traditional risk factors predict severe coronavirus disease 2019 (COVID-19) is uncertain, in part, because of potential confounding by age and sex.

**Methods:** We performed a systematic review of studies that explored pre-existing CVD and its traditional risk factors as risk factors of severe COVID-19 (defined as death, acute respiratory distress syndrome, mechanical ventilation, or intensive care unit admission). We searched PubMed and Embase for papers in English with original data (≥10 cases of severe COVID-19). Using random-effects models, we pooled relative risk (RR) estimates and conducted meta-regression analyses.

**Results:** Of the 661 publications identified in our search, 25 papers met our inclusion criteria, with 76,638 COVID-19 patients including 11,766 severe cases. Older age was consistently associated with severe COVID-19 in all eight eligible studies, with RR >∼5 in >60-65 vs. <50 years. Three studies showed no change in the RR of age after adjusting for covariate(s). In univariate analyses, factors robustly associated with severe COVID-19 were male sex (10 studies; pooled RR=1.73, [95%CI 1.50-2.01]), hypertension (8 studies; 2.87 [2.09-3.93]), diabetes (9 studies; 3.20 [2.26-4.53]), and CVD (10 studies; 4.97 [3.76-6.58]). RR for male sex was likely to be independent of age. For the other three factors, meta-regression analyses suggested confounding by age. Only four studies reported multivariable analysis, but most of them showed adjusted RR ∼2 for hypertension, diabetes, and CVD. No study explored renin-angiotensin system inhibitors as a risk factor for severe COVID-19.

**Conclusions:** Despite the potential for confounding, these results suggest that hypertension, diabetes, and CVD are independently associated with severe COVID-19 and, together with age and male sex, can be used to inform objective decisions on COVID-19 testing, clinical management, and workforce planning.

## Introduction

Cases of coronavirus disease 2019 (COVID-19) are rapidly increasing globally. As of April 25, 2020, more than 2.8 million cases have been confirmed and ∼200,000 deaths have been reported in ∼190 countries.^1^ Several studies have rapidly provided crucial data (e.g., incubation period) related to various aspects of the novel coronavirus (SARS-CoV-2: severe acute respiratory syndrome coronavirus 2) infection.^2^ However, risk factors for the severity and prognosis of COVID-19 are poorly understood. Such information is critical to identify high risk patients and to facilitate planning (e.g., forecasting the need for hospital beds and mechanical ventilators). These risk factors also have implications for workforce allocation (e.g., assignment of healthcare providers with specific risk factors to positions with reduced risk of exposure to COVID-19).

To date, several studies have reported that a history of cardiovascular disease (CVD) and traditional CVD risk factors, e.g., age, male sex, current smoking, hypertension, and diabetes, are associated with severe COVID-19. However, other than age, results have been inconsistent. Furthermore, few studies that evaluated other potential risk factors accounted for potential confounding by age and sex. For example, some studies reported that hypertension is a risk factor of severe COVID-19, but their observations may simply reflect the fact that hypertension is more common in older adults.^3^ Nonetheless, despite the lack of robust evidence, this observation, together with the fact SARS-CoV-2 uses angiotensin-converting enzyme 2 as an entry to human cell,^4^ has raised a concern about continued use of renin-angiotensin system increase among some clinicians and researchers.^5,6^

In this context, we conducted a systematic review of studies reporting cardiovascular risk factors and their relation to severe manifestation of COVID-19 (i.e., death, acute respiratory distress syndrome [ARDS], the need of mechanical ventilator support, and admission to an intensive care unit [ICU]), with a particular interest on studies that adjusted for key confounders such as age and sex.

## Methods

### Search strategy

We conducted this systematic review following the PRISMA Statement. According to the pre-determined protocol, we systematically searched PubMed and Embase for eligible reports (search terms are listed in Web Appendix 1). We included full reports or letters with original data written in English. Eligible study designs were cohort study, cross-sectional, case series, case-control study, and clinical trials. We conducted the literature search on April 3, 2020 and restricted to publications after December 1, 2019.

Our review included studies that reported adult patients, aged 18 years or older. There was no restriction with respect to gender, race/ethnicity, and comorbidities. The primary outcome of interest was severe COVID-19 defined by any of the following: all-cause mortality, ICU admission, ARDS, or the need for mechanical ventilation. We included studies reporting at least 10 cases of severe COVID-19. To obtain reliable estimates with enough number of outcomes and considering clinical cascade (e.g., death as the final outcome), when one study reported results for multiple outcomes, we prioritized any composite outcome followed by ICU admission, ARDS, the need of mechanical ventilation, and mortality.

Potential risk factors of interest were pre-existing CVD (including cardiac disease and cerebrovascular disease) and its traditional risk factors recognized in major CVD clinical guidelines: age, sex, smoking, hypertension, dyslipidemia, and diabetes.^7^ We found only one study reporting severity of COVID-19 by dyslipidemia (low-density lipoprotein). We categorized risk factors into sociodemographic factors (age, sex, and smoking) and clinical factors (hypertension, diabetes, and pre-existing CVD).

### Study selection

One author (N.D.) conducted the literature search, exported eligible publications to EndNote X8 reference manager (Thomson Reuters, Philadelphia, Pennsylvania), and uploaded them to Covidence (Melbourne, Australia), a platform for literature screening. Eight reviewers (N.D., M.K., Y.G., Y.H., Y.M., X.H., M.C., and J.I.) worked in pairs to independently screen titles and abstracts. For research letters without abstracts, reviewers used text content for the initial screening. All conflicts were resolved by one of two lead reviewers (N.D., M.K.). For all publications accepted at this step, the same pairs performed the full detailed text review to evaluate final eligibility. The two lead reviewers resolved any conflicts.

### Data collection and quality assessment

The same eight reviewers collected relevant data elements from each identified publication and recorded in Microsoft Excel (Microsoft Corporation, Redmond, Washington, USA). Overall quality was based on the Newcastle Ottawa Quality Assessment Scale (NOS),^8^ which includes eight items about selection, comparability, and outcome (Web Appendix 2). The NOS score for cohorts studies ranges from 0 to 9; a score greater than 6 was considered high-quality. For cross-sectional and case-control studies, we applied an adapted form of the NOS.^9^ The maximum score was 10 and 9, and 7 and 6 points were used to identify studies with high quality, respectively. The paired reviewers resolved conflicts related to their own data collection and quality assessment.

### Data synthesis and analysis

We summarized relative risk estimates (odds ratios or hazard ratios) of the association between each risk factor and the primary outcome from the relevant studies. We pooled these estimates using random-effects meta-analysis. When studies did not report these measures of association but the prevalence of risk factors of interest by the outcome status (e.g., survivors vs. non-survivors), we calculated crude odds ratios and their 95% CIs. In this process, when there was any cell with zero count, we added 0.5 to each cell, as appropriate.^10^

Potential confounding by age and sex is relevant to prior CVD, hypertension, and diabetes, since these comorbidities become more prevalent with increased age.^3^ Because most studies did not report adjusted risk estimates for these comorbidities, we conducted meta-regression analyses with random-effects for log odds ratio or log hazard ratio for these comorbidities by the difference in mean or median age between those with vs. without primary outcome across eligible studies. To obtain reliable estimates, we required at least five studies for each meta-regression analysis. We also depicted funnel plots and visually checked the possibility of publication bias. Heterogeneity of study estimates was assessed by *I*^2^ statistic, and *I*^2^ >75% was considered high heterogeneity.^11^

In our review, we found a total of 10 reports likely using the same data source even though they did not analyze identical samples. Since we could not be certain whether these were duplicate or overlapping publications, we had two approaches to conducting the meta-analyses: (1) a restrictive approach in which we excluded reports with high possibility of overlap [e.g., data from the same hospitals during the same period] and (2) an inclusive approach in which we included all publications. For the restrictive approach, we used the largest study with estimates of interest. To be conservative, we present the restrictive meta-analysis as our primary analysis. The results of the inclusive meta-analysis are available in the supplemental material. A p-value <0.05 was considered statistically significant. All analyses were conducted with Stata 14 or 15 (StataCorp, LLC, College Station, Texas, USA).

## Results

### Search results

Our systematic review identified 661 potentially eligible publications after removing duplicate publications (Figure 1). Of these, 585 publications were excluded after screening titles and abstracts. Of the remaining 76 publications reviewed with full-text screening, we excluded 51 publications that did not meet our inclusion criteria, leaving 25 publications^12-36^ for analyses. Most of these publications were considered high quality (Web Table 1). Of the included studies, death was reported in 21 studies, ICU admission in 11 studies, ARDS in 13 studies, and mechanical ventilation in 11 studies. Three studies reported a composite outcome.

**Table 1.** Summary of 25 studies included in systematic review

| Reference | Journal | Region | Setting | Sample size | Severe, % | Age median (IQR) or mean (SD) in y | Male, % | Current smoking, % | HTN, % | DM, % | CVD, % |
| --- | --- | --- | --- | --- | --- | --- | --- | --- | --- | --- | --- |
| Bhatraju PK | NEJM | USA | 9 Seattle-area hospitals | 24 | 75 | 64 (18) | 63 | 22* | - | 58 | 8 |
| Cao JL | Intensive Care Med | China | Zhongnan Hospital | 102 | 17.6 | 54 (37-67) | 52 | - | 27.5 | 10.8 | 4.9 |
| Chen J | J Infect | China | Shanghai Public Health Clinical Center | 249 | 8.8 | 51 (36–64) | 50.6 | - | - | - | 21.7 |
| Chen T | Bmj | China | Tongji Hospital | 274 | 72 | 62 (44-70) | 62 | 4 | 34 | 17 | 8 |
| Cheng YC | Kidney International | China | Tongji Hospital | 701 | 16.1 | 63 (50-71) | 52.4 | - | 33.4 | 14.3 |  |
| China CDC | CCDC | China | China National Data | 44,672 | 18.5 | - | 51.4 | - | 12.8 | 5.3 | 4.2 |
| Deng Y | Chin Med J | China | Tongji Hospital and Central Hospital of Wuhan | 225 | 48 | - | 55 | - | 26 | 12 | 8 |
| Guan W | NEJM | China | 552 hospitals in 30 provinces | 1099 | 6.5 | 47 (35-58) | 58.1 | 12.6 | 15 | 7.4 | 2.5 |
| Guo T | JAMA Cardiol | China | Seventh Hospital of Wuhan | 187 | 23 | 59 (15) | 49 | 10 | 33 | 15 | 11 |
| Huang CL | Lancet | China | Jinyintan Hospital | 41 | 32 | 49 (41-58) | 73 | 7 | - | - | 15 |
| Lian JS | Clin Infect Dis | China | Hospitals in Zhejiang province | 788 | 9.8 | 45.83 (14.87) | 51.6 | 6.9 | 16 | 7.2 | 1.4 |
| Liang WH | Lancet Oncology | China | 575 hospitals across China | 1590 | 8.2 | 48.8±16.2 | 57.2 | 7 | - | - | - |
| Onder G | JAMA | Italy | Italy National Data | 22512 | 7.2 | - | - | - | - | - | - |
| Ruan QR | Intensive Care Med | China | Jinyintan Hospital and Tongji Hospital | 150 | 45 | - | 68 | - | 35 | - | 8.7 |
| Shi SB | JAMA Cardiol | China | Zhejiang Province | 416 | 23.3 | 64 (21-95) | 49.3 | - | 30.5 | 14.4 | 10.6 |
| Tang N | J Thromb Haemost | China | Tongji Hospital | 183 | 11.5 | 54.1±16.2 | 53.6 | - | - | - | - |
| US CDC | MMWR | US | US National Data | 2449 | 4.9 | - | - | - | - | - | - |
| US CDC_2 | MMWR | USA | US National Data | 6637 | 6.4 | - | - | 1.3 | 1.6 | 10.9 | 9.0 |
| Wang DW | JAMA | China | Zhongnan Hospital | 138 | 26.1 | 56 (42-68) | 54.3 | - | 31.2 | 10.1 | 14.5 |
| Wang L | J Infect Mar | China | Renmin Hospital of Wuhan University | 339 | 23.6 | 69 (65-76) | 49 | - | 40.8 | 16 | 15.7 |
| Wu CM | JAMA Intern Med | China | Jinyintan Hospital, Wuhan | 201 | 41.8 | 51 (43-60) | 63.7 | - | 19.4 | 10.9 | 4 |
| Yang XB | Lancet Respir | China | Jinyintan Hospital, Wuhan | 52 | 67 | 59.7 (13.3) | 67 | 4 | - | 17 | 10 |
| Yuan ML | PLoS One | China | Central Hospital of Wuhan | 27 | 41 | 60 (47-69) | 45 | - | 19 | 22 | 11 |
| Zhang L | Ann Oncol | China | Tongji Sino-French New Town Hospital, Union Red Cross Hospital and Union West Hospital | 28 | 53.6 | 65 (56-70) | 60.7 | - | - | 14.3 | 14.3 |
| Zhou F | Lancet | China | Jinyintan Hospital and Wuhan Pulmonary Hospital | 191 | 31 | 56 (46-67) | 62 | 6 | 30 | 19 | 8 |

All studies were published in 2020.
Abbreviations: COVID-19: coronavirus disease 2019; CVD: cardiovascular disease; DM: diabetes; HTN: hypertension; NR, not reported.
\*Current or former smoking

**Figure 1.**
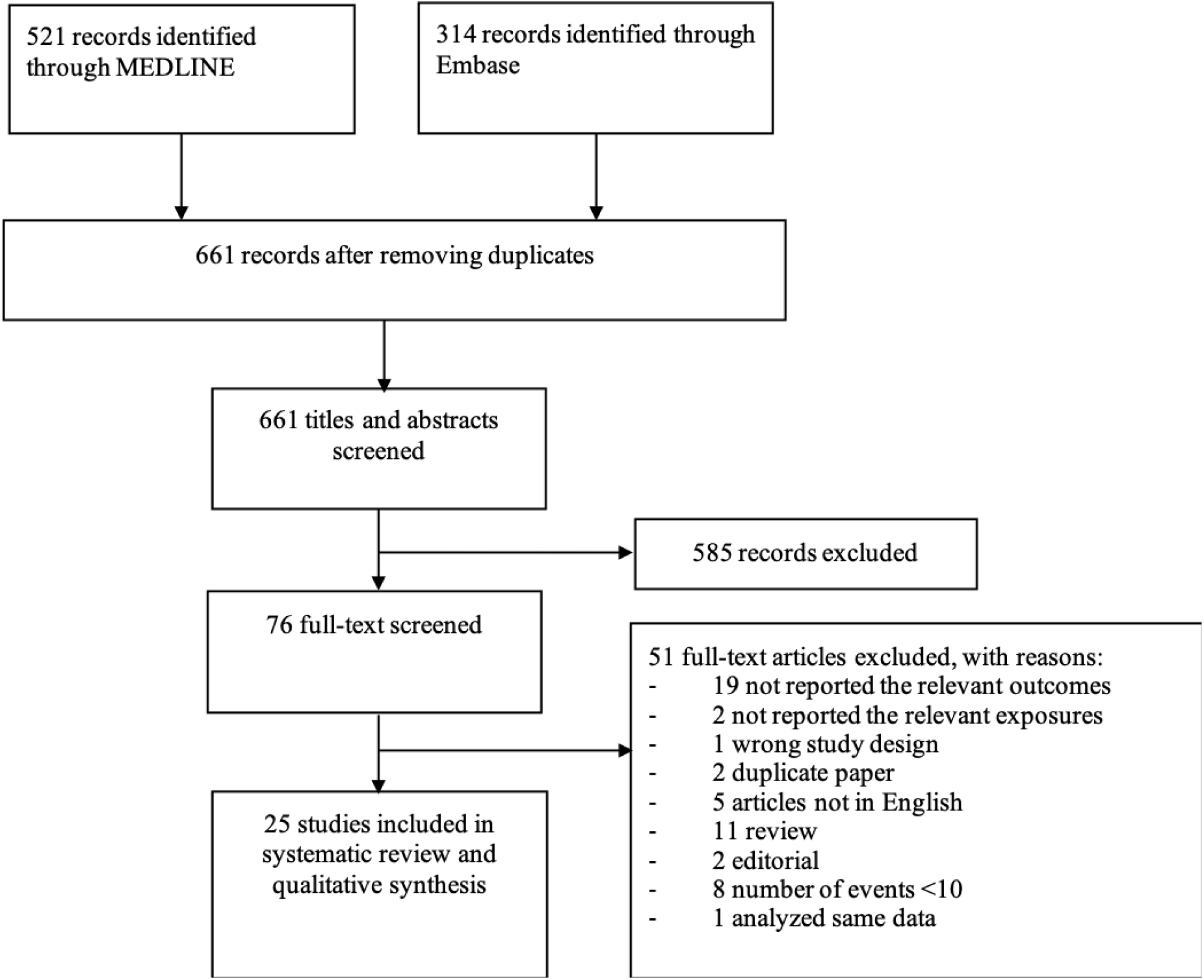
Flow chart of study selection

### Study characteristics

Most studies reported COVID-19 patients from China (21 studies) and were small with sample size <300 (15 studies) (Table 1). All studies included confirmed COVID-19 patients with laboratory tests. A total of 76,638 COVID-19 patients were included in these studies, with 11,766 patients manifesting severe disease. Most studies exclusively investigated hospitalized COVID-19 cases. Average or median age ranged from 46 to 69 years, and 45% to 73% of participants were male. Most studies reported the prevalence of CVD risk factors; the prevalence of hypertension ranged from 1.6% to 40.8%, diabetes from 5.3% to 58%, and pre-existing CVD from 1.4% to 21.7%.

### Sociodemographic factors: age, sex, and smoking

Older age was associated with higher risk of severe COVID-19 in all 16 studies with relevant estimates (Table 2). This pattern was confirmed in national surveys database in the US, China, and Italy, without any clear thresholds (Web Table 2). The case fatality rate in those three databases exceeded 1% around age of 50-55 years and 10% above 80-85 years (above 70 years in Italy). Three studies reported the relative risk by age in both unadjusted and multivariable models and did not observe material changes in the effect size of age after adjustment for comorbidities.^14,35^

**Table 2.**
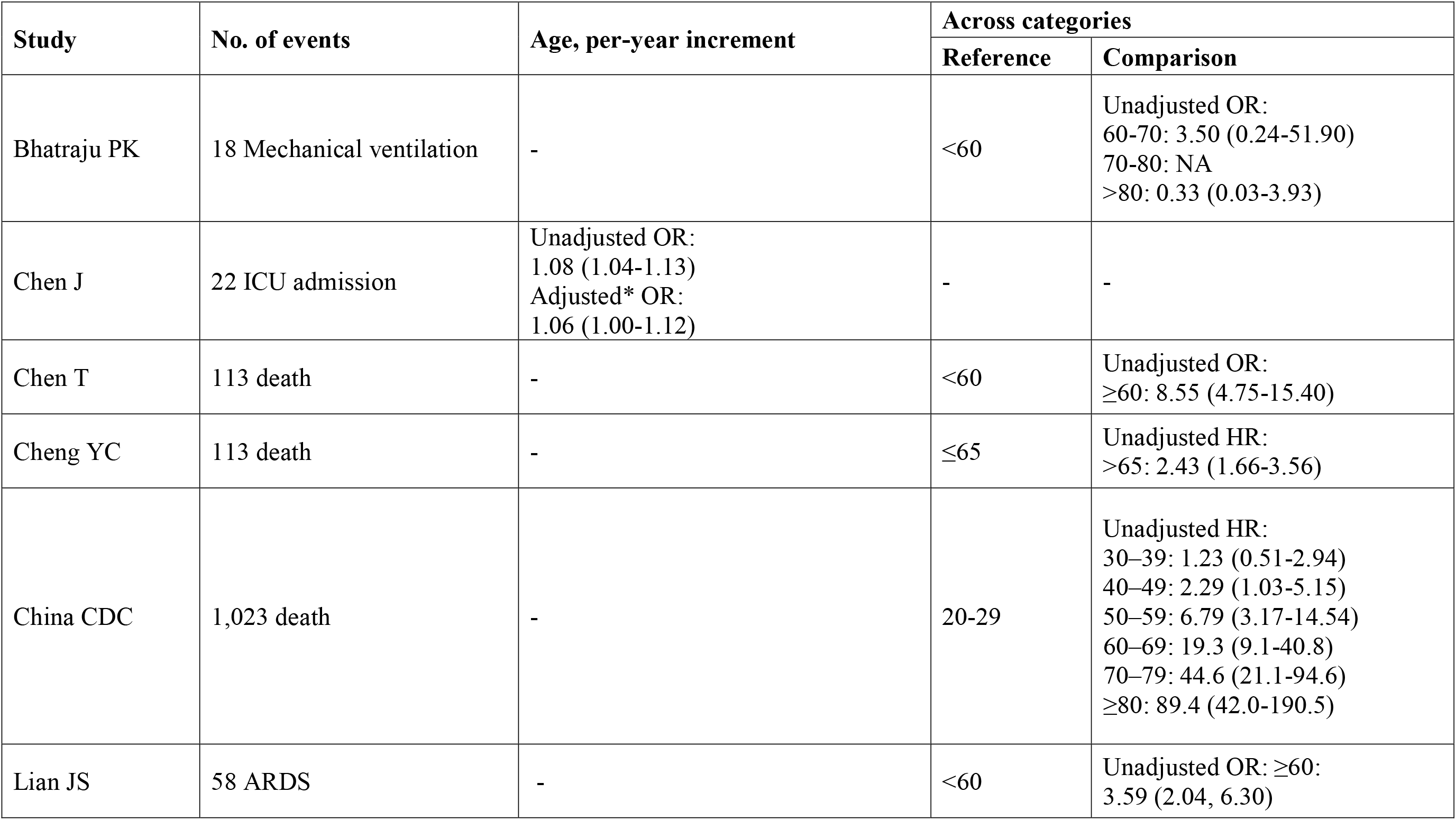
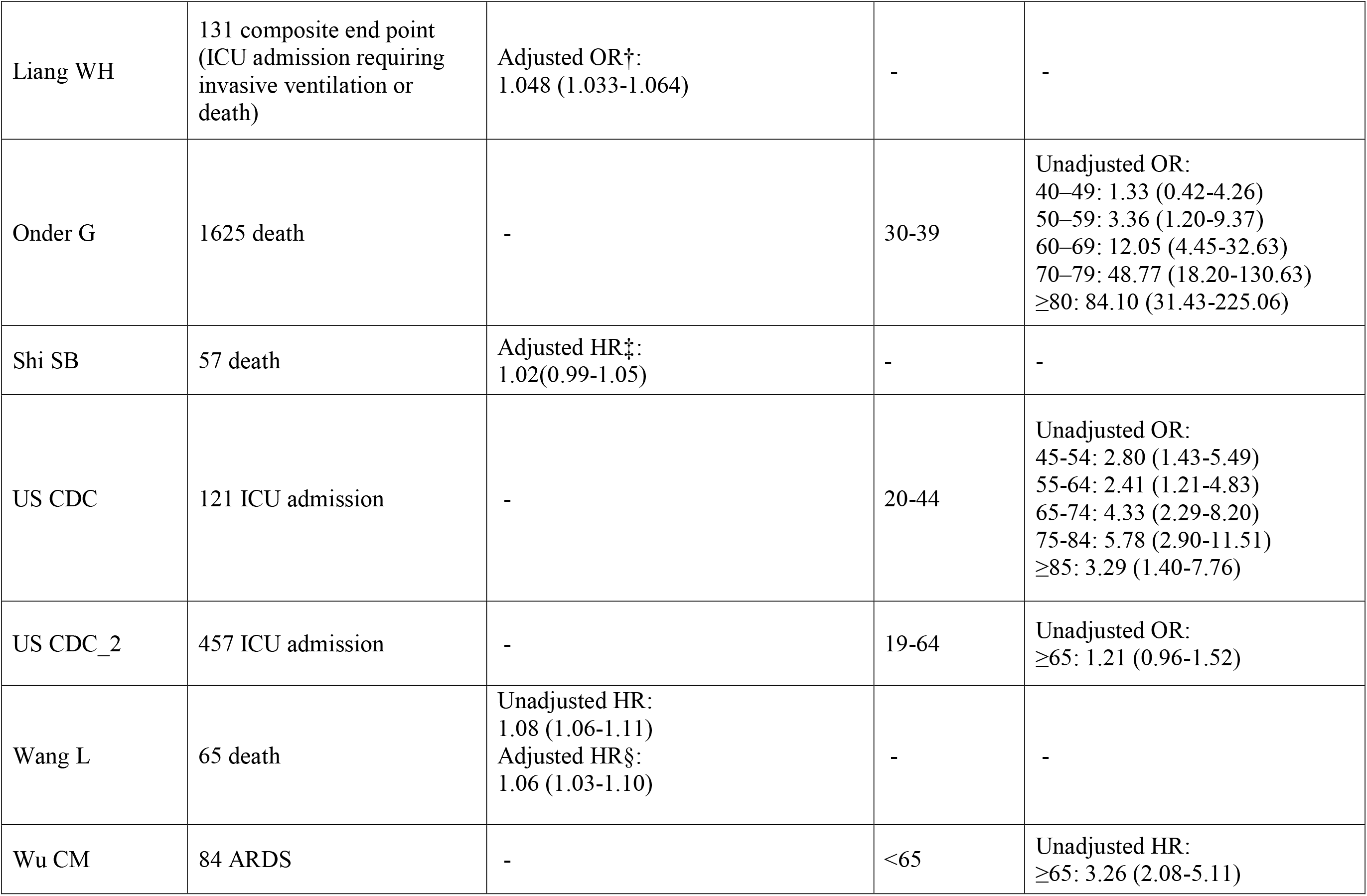
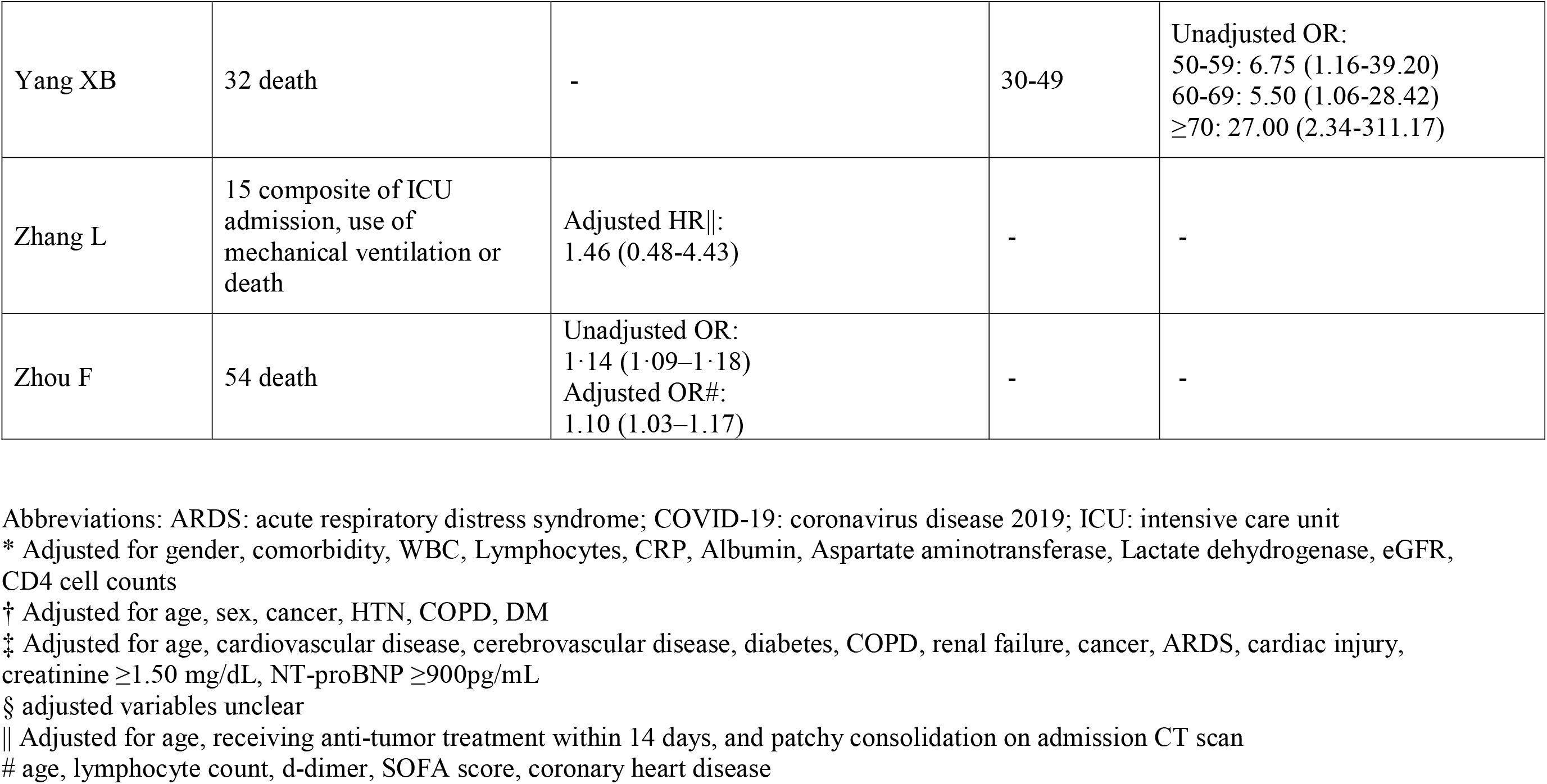
Relative risk estimates of severe COVID-19 by age in eligible studies

| Study | No. of events | Age, per-year increment | Across categories |  |
| --- | --- | --- | --- | --- |
|  |  |  | Reference | Comparison |
| Bhatraju PK | 18 Mechanical ventilation | - | <60 | Unadjusted OR:<br>60-70: 3.50 (0.24-51.90)<br>70-80: NA<br>>80: 0.33 (0.03-3.93) |
| Chen J | 22 ICU admission | Unadjusted OR:<br>1.08 (1.04-1.13)<br>Adjusted* OR:<br>1.06 (1.00-1.12) | - | - |
| Chen T | 113 death | - | <60 | Unadjusted OR:<br>≥60: 8.55 (4.75-15.40) |
| Cheng YC | 113 death | - | ≤65 | Unadjusted HR:<br>>65: 2.43 (1.66-3.56) |
| China CDC | 1,023 death | - | 20-29 | Unadjusted HR:<br>30-39: 1.23 (0.51-2.94)<br>40-49: 2.29 (1.03-5.15)<br>50-59: 6.79 (3.17-14.54)<br>60-69: 19.3 (9.1-40.8)<br>70-79: 44.6 (21.1-94.6)<br>≥80: 89.4 (42.0-190.5) |
| Lian JS | 58 ARDS | - | <60 | Unadjusted OR: ≥60:<br>3.59 (2.04, 6.30) |
| Liang WH | 131 composite end point<br>(ICU admission requiring<br>invasive ventilation or<br>death) | Adjusted OR†:<br>1.048 (1.033-1.064) | - | - |
| Onder G | 1625 death | - | 30-39 | Unadjusted OR:<br>40-49: 1.33 (0.42-4.26)<br>50-59: 3.36 (1.20-9.37)<br>60-69: 12.05 (4.45-32.63)<br>70-79: 48.77 (18.20-130.63)<br>≥80: 84.10 (31.43-225.06) |
| Shi SB | 57 death | Adjusted HR‡:<br>1.02(0.99-1.05) | - | - |
| US CDC | 121 ICU admission | - | 20-44 | Unadjusted OR:<br>45-54: 2.80 (1.43-5.49)<br>55-64: 2.41 (1.21-4.83)<br>65-74: 4.33 (2.29-8.20)<br>75-84: 5.78 (2.90-11.51)<br>≥85: 3.29 (1.40-7.76) |
| US CDC_2 | 457 ICU admission | - | 19-64 | Unadjusted OR:<br>≥65: 1.21 (0.96-1.52) |
| Wang L | 65 death | Unadjusted HR:<br>1.08 (1.06-1.11)<br>Adjusted HR§:<br>1.06 (1.03-1.10) | - | - |
| Wu CM | 84 ARDS | - | <65 | Unadjusted HR:<br>≥65: 3.26 (2.08-5.11) |
| Yang XB | 32 death | - | 30-49 | Unadjusted OR:<br>50-59: 6.75 (1.16-39.20)<br>60-69: 5.50 (1.06-28.42)<br>≥70: 27.00 (2.34-311.17) |
| Zhang L | 15 composite of ICU admission, use of mechanical ventilation or death | Adjusted HR\|\|:<br>1.46 (0.48-4.43) | - | - |
| Zhou F | 54 death | Unadjusted OR:<br>1.14 (1.09–1.18)<br>Adjusted OR#:<br>1.10 (1.03–1.17) | - | - |

Abbreviations: ARDS: acute respiratory distress syndrome; COVID-19: coronavirus disease 2019; ICU: intensive care unit
\* Adjusted for gender, comorbidity, WBC, Lymphocytes, CRP, Albumin, Aspartate aminotransferase, Lactate dehydrogenase, eGFR, CD4 cell counts
† Adjusted for age, sex, cancer, HTN, COPD, DM
‡ Adjusted for age, cardiovascular disease, cerebrovascular disease, diabetes, COPD, renal failure, cancer, ARDS, cardiac injury, creatinine ≥1.50 mg/dL, NT-proBNP ≥900pg/mL
§ adjusted variables unclear
|| Adjusted for age, receiving anti-tumor treatment within 14 days, and patchy consolidation on admission CT scan

Most studies showed a higher risk of severe COVID-19 in men than in women, with a pooled crude relative risk estimate of severe COVID-19 between men and women of 1.73 (95% CI 1.50-2.01) (Figure 2A and Web Figure 1A). In meta-regression analyses, there was no substantial evidence of confounding by age (Web Figure 2). A funnel plot did not indicate major publication bias (Web Figure 3A).

**Figure 2.**
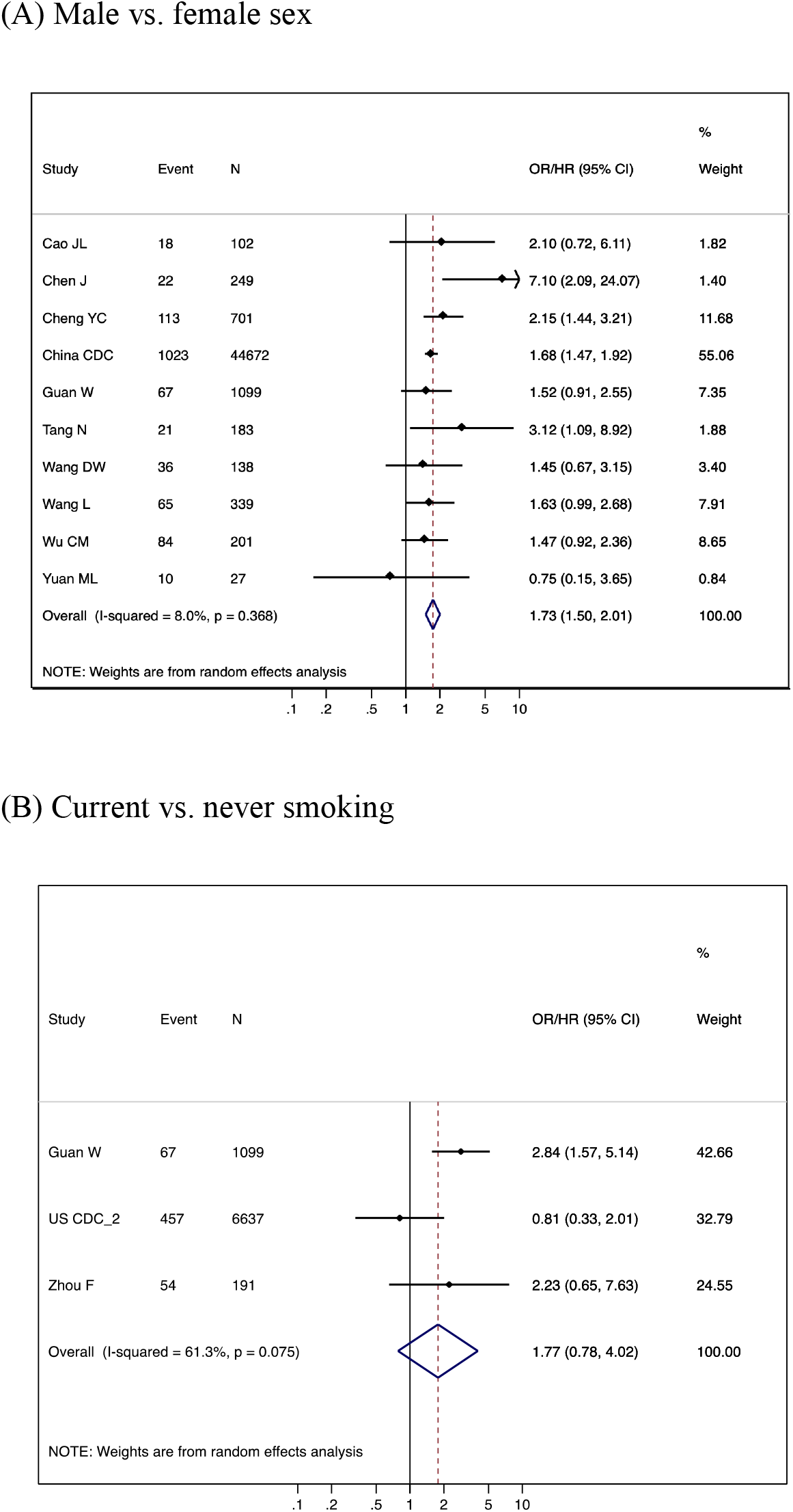
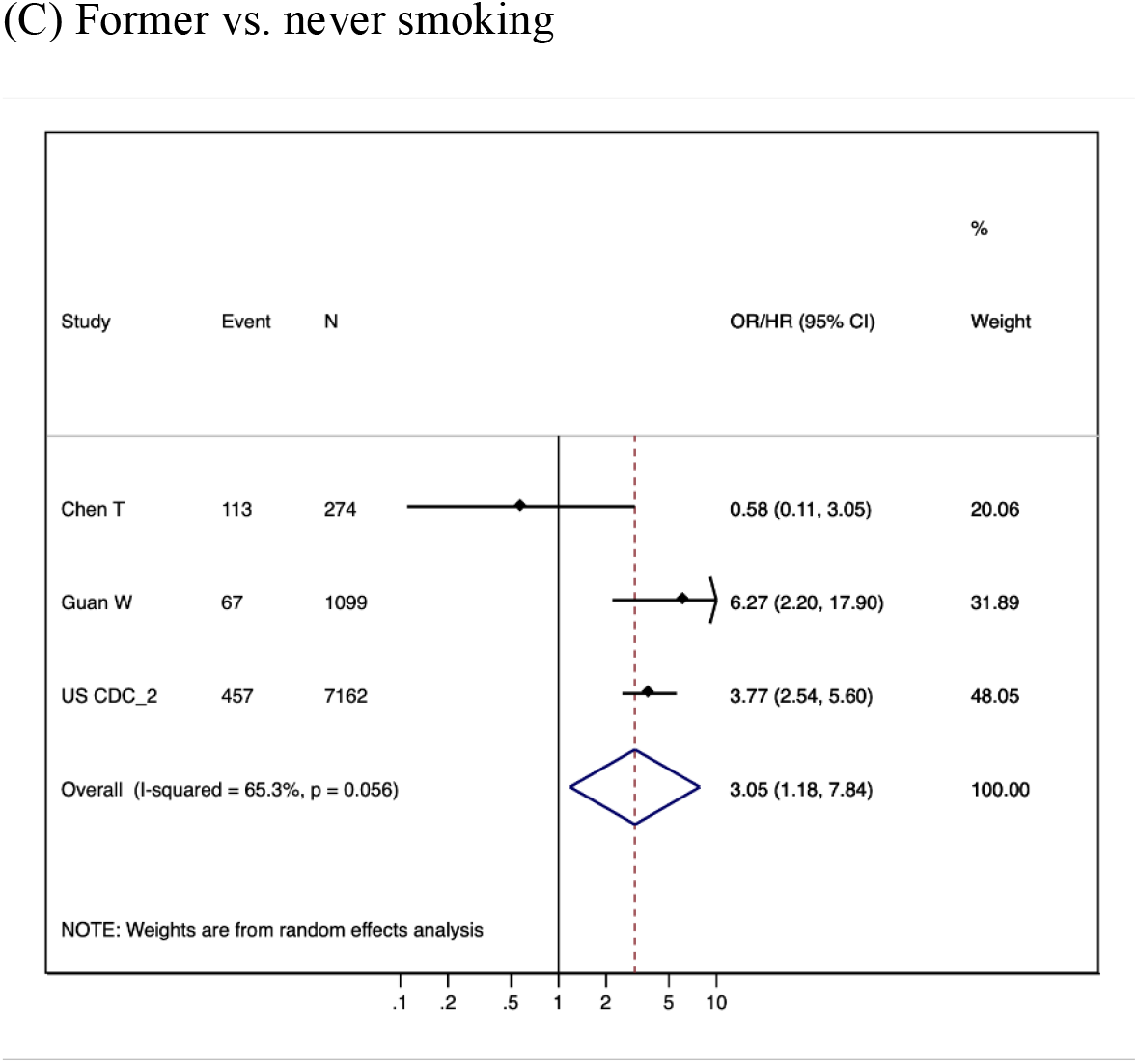
Forest plots of unadjusted relative risk estimates of severe COVID-19 according to gender (A), smoking status (B and C) based on restrictive meta-analysis

**Figure 3.**
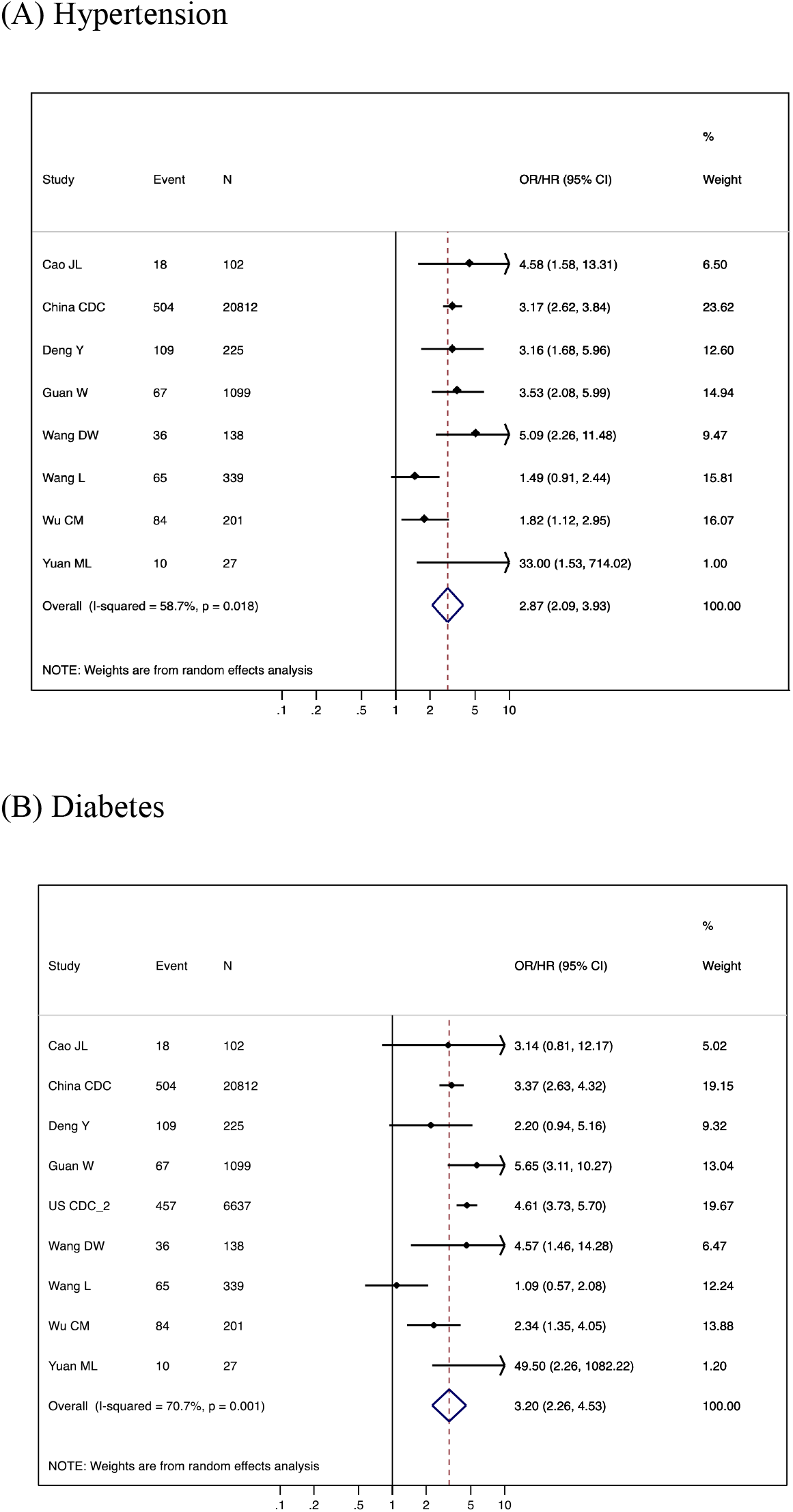
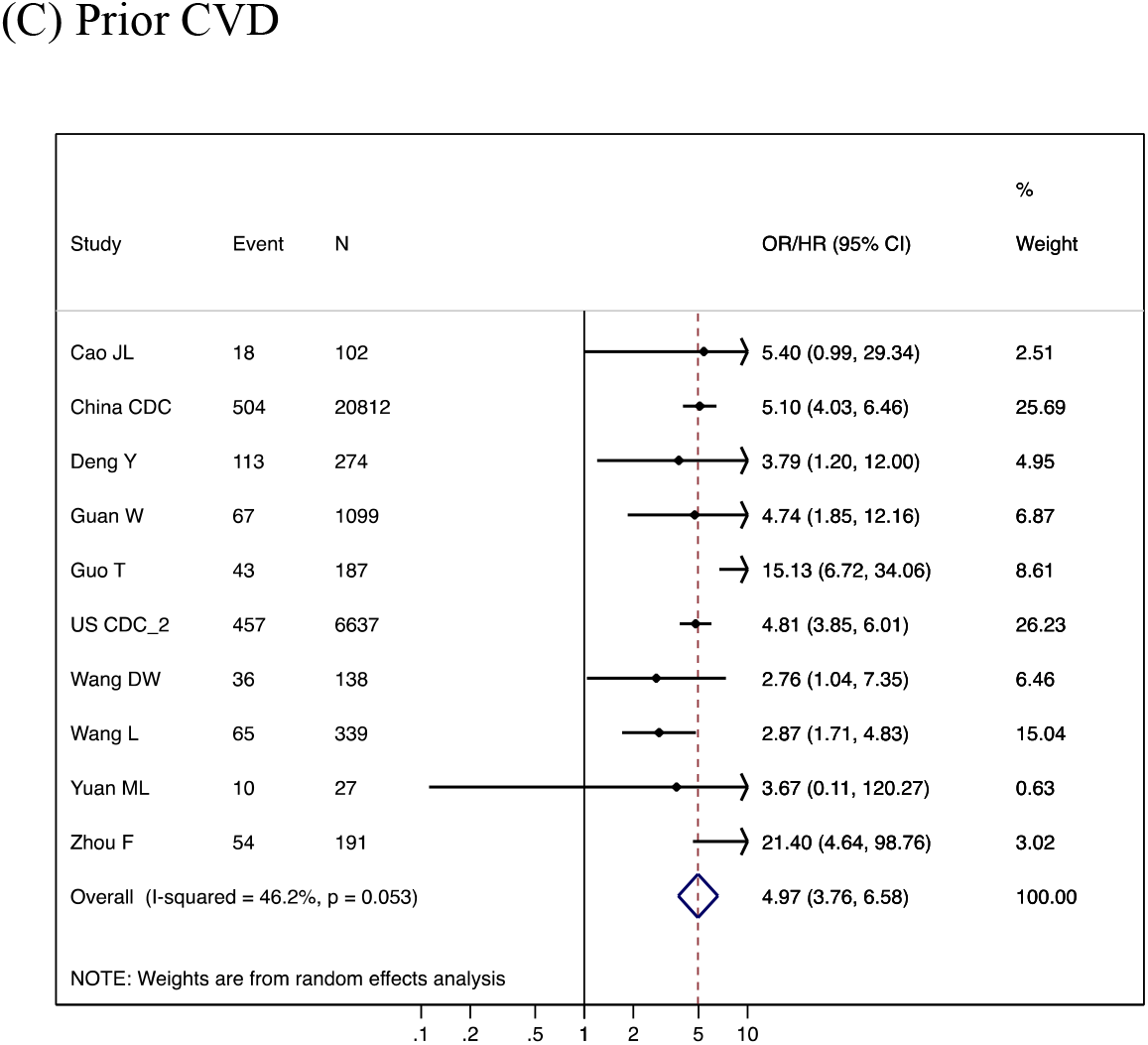
Forest plots of unadjusted relative risk estimates of severe COVID-19 according to hypertension (A), diabetes (B), and prior CVD (C) on restrictive meta-analysis The event number and total number of China CDC paper are different from Tables 1 and 2 because of missing value in comorbidity.

Only a couple of studies reported associations of current smoking with severe COVID-19, with only one study reaching statistical significance (Figure 2B and Web Figure 1B). The pooled estimate of relative risk for severe COVID-19 was ∼1.8 in both restrictive and inclusive meta-analyses, although statistical significance was restricted to the latter. Three studies reported the association between former vs. never smoking, with the pooled relative risk estimates of 3.05 (1.18-7.84) (Figure 2C). The corresponding funnel plot is shown in Web Figure 3B.

### Clinical factors: hypertension, diabetes, and prior CVD

All eligible studies with data on hypertension reported a positive association of hypertension with severe COVID-19 (Figure 3A and Web Figure 4A), with the primary pooled relative risk estimate of 2.87 (95% CI 2.09-3.93). Most eligible studies also demonstrated a positive association between diabetes and severe COVID-19 (Figure 3B and Web Figure 4B), with the primary pooled relative risk estimate of 3.20 (2.26-4.53). Similarly, a majority of eligible studies showed a positive association between prior CVD and severe COVID-19, with the primary pooled relative risk of 4.97 (3.76-6.58) (Figure 3C and Web Figure 4C). None of these meta-analyses demonstrated high heterogeneity with *I*^2^ >75%. There was no indication of publication bias for these comorbidities (Web Figure 3C-E). In the one available study that assessed low-density lipoprotein as a risk factor of COVID-19, Wu et al reported that higher levels were associated with lower risk of severe COVID-19 (0.63 [0.44-0.88] per 1 mmol/L increment)^32^; no other lipid fractions were reported. No study explored renin-angiotensin system inhibitors as a risk factor for severe COVID-19.

**Figure 4.**
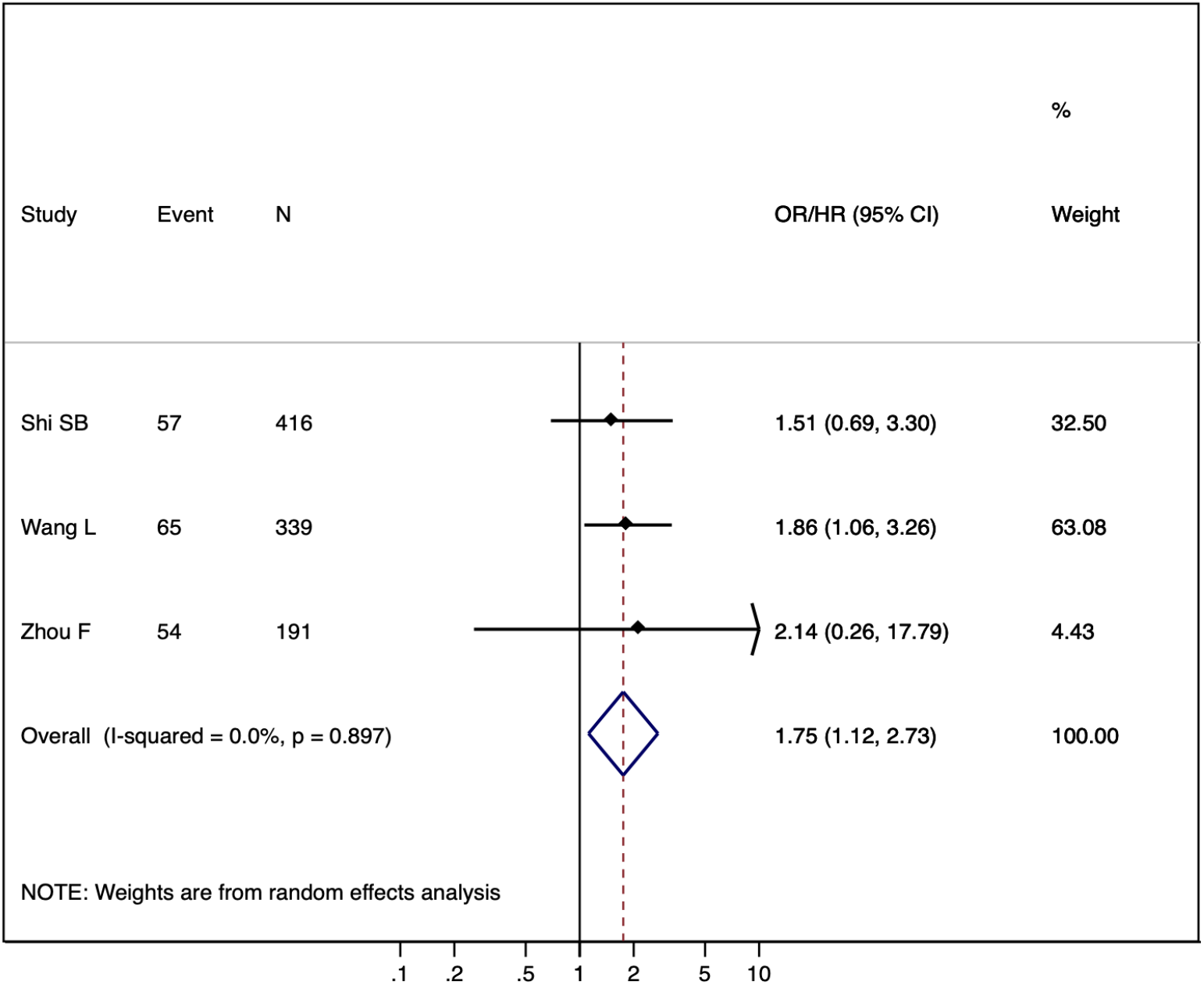
Forest plots of adjusted relative risk estimates of severe COVID-19 according to prior CVD

### Potential confounding by age and sex

Among the identified studies, only four studies reported adjusted relative risk estimates of these clinical factors. For pre-existing CVD, the pooled adjusted relative risk estimate across three studies was 1.75 (1.12-2.73) (Figure 4). Although these studies accounted for different confounders, all included age. Two studies reported adjusted relative risk for diabetes, but one of them was likely to have overadjustment bias by including potential mediators such as kidney failure, ARDS, and end organ damage markers (natriuretic peptide, cardiac troponin, and serum creatinine). The other study by Liang et al with 1,590 patients reported an adjusted odds ratio of 2.21 (1.33-3.66) for diabetes.^23^ This study also showed the adjusted odds ratio of 1.88 (1.22-2.90) for hypertension.

Meta-regression analyses demonstrated that studies with greater age difference between those with vs. without severe COVID-19 tended to have greater relative risk according to the presence of hypertension, diabetes, and pre-existing CVD, indicating some levels of potential confounding by age (Figure 5 and Web Figure 5), although the statistical significance was only seen for hypertension in the inclusive meta-analysis (Web Figure 5A). The meta-regression analysis did not indicate that a higher proportion of male sex confounded the association of these clinical factors with severe COVID-19 (Web Figure 6).

**Figure 5.**
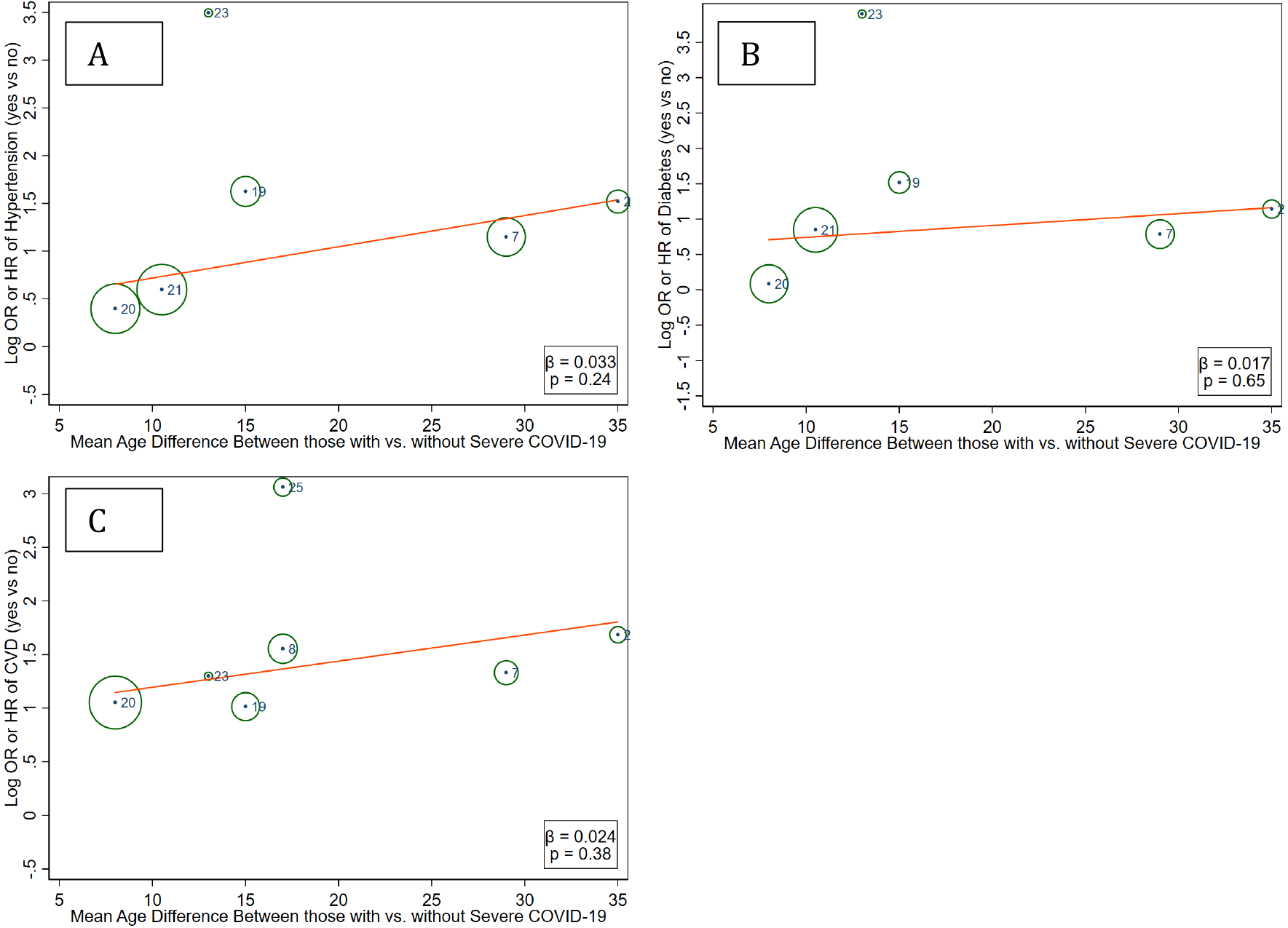
Meta-regression of unadjusted relative risk of severe COVID-19 for hypertension (A), diabetes (B), and CVD (C) by age difference between severe vs. non-severe COVID-19 based on restrictive meta-analysis List of studies: 1 Bhatraju PK et al, N Engl J Med; 2 Cao JL et al, Intensive Care Med; 3 Chen J et al, J Infect; 4 Chen T et al, BMJ; 5 Cheng YC et al, Kidney International; 6 China CDC, CCDC Weekly; 7 Deng Y et al, Chin Med J (Engl); 8 Guan W et al, N Eng J Med; 9 Guo T et al, JAMA Cardiol; 10 Huang CL et al, Lancet; 11 Lian JS et al, The Lancet Oncology; 12 Liang WH et al, The Lancet Oncology; 13 Onder G et al, JAMA; 14 Ruan QR et al, Intensive Care Med; 15 Shi SB et al, JAMA Cardiol; 16 Tang et al, J Thromb Haemost; 17 US CDC, MMWR; 18 US CDC_2, MMWR; 19 Wang DW et al, JAMA; 20 Wang L et al, J Infect Mar; 21 Wu CM et al, JAMA Intern Med; 22 Yang XB et al, Lancet Respir; 23 Yuan ML et al, PLoS One; 24 Zhang L et al, Ann Oncol; 25 Zhou F et al, Lancet

## Discussion

To our knowledge, this is the first systematic review and meta-analysis focusing on the relationship of severe COVID-19 with CVD and its risk factors. We confirmed a robust association of age and male sex with severe COVID-19. Their contributions are likely to be independent of each other. A few studies demonstrated positive associations of current and former smoking with severe COVID-19. Several studies reported that pre-existing CVD, hypertension, and diabetes were also associated with severe COVID-19. However, only four studies reported estimates for these comorbidities adjusted for age and/or sex. Across those studies, we could only meaningfully pool estimates for pre-existing CVD, which demonstrated a significant relative risk of ∼1.8. One relatively large study by Liang et al showed independent associations of hypertension and diabetes with severe COVID-19 in analyses that adjusted for age and a few other comorbidities.^23^ Although the primary estimate was not statistically significant, our meta-regression analyses indicated some degree of confounding by age, but not necessarily by sex, for the associations of hypertension, diabetes, and prior CVD with severe COVID-19.

The positive association of sociodemographic factors (age, male sex, and smoking) with severe COVID-19 is consistent with reports of other infectious diseases (e.g., influenza virus and SARS in 2003).^37-39^ There are several plausible mechanisms. Older age is linked to reduced immune reaction, more comorbidities, and limited organ reserve.^3,37,40^ Male sex is related to higher prevalence of comorbidities, less frequency of washing hands, and immunological disadvantage given X-chromosome coding proteins in the immune system,^2,41,42^ whereas smoking can damage respiratory system.^43^

In our meta-analysis, we confirmed overall positive crude associations of CVD, hypertension, and diabetes with severe COVID-19, with pooled relative risk estimates around 3-5. An important question is whether these associations are independent of major confounders, particularly age. In this regard, the meta-analysis of three studies indicated an independent association for pre-existing CVD. For hypertension and diabetes, we could not obtain a definite answer as very few studies ran multivariable models, although our meta-regression analysis indicated some degree of confounding by age. Of note, Liang et al showed independent associations of hypertension and diabetes with severe COVID-19 in a relatively large sample size of 1,590 patients. These is also biologic plausibility. Hypertension and diabetes are leading risk factors for CVD and kidney diseases, and there is evidence that COVID-19 damages these organs.^26,44^

However, we should not conflate the observed associations of hypertension and diabetes with severity of COVID-19 infection as indicative of adverse effects of renin-angiotensin system inhibitors. None of the 25 studies in our systematic review explored these medications regarding severe COVID-19. Moreover, in the only study reporting the prevalence of renin-angiotensin system inhibitors use,^16^ ∼30% of patients reported prevalent hypertension, but only ∼5% of patients were taking renin-angiotensin system inhibitors.^16^ Thus, it is reasonable that many expert organizations recommend continuing this category of medications until better evidence becomes available.^4^

Our results can potentially be used to guide decision-making. While the lack of discrete age thresholds for severe COVID-19 complicates this process, data on case fatality rates from three countries (Web Table 2) suggest that age older than 60 or 65 years confers high risk of severe COVID-19 (relative risk > ∼5 compared to <50 years). Our results also indicate male sex is an independent risk factor with a pooled relative risk of ∼1.7. Although the pooled crude relative risk of ∼3-5 for hypertension, diabetes, and CVD probably overestimate their impact beyond age and sex, all or some of them are likely to each confer 1.5-2 times greater risk. Thus, using these factors, it is possible to estimate, at least crudely, the risk of severe COVID-19. For example, a man aged 60-65 years old, with either hypertension, diabetes, or prior CVD would have a risk of ∼15 fold higher risk (approximation of 5 × 1.7 × 2) compared to a woman younger than 50 years without any of these clinical factors. Such information could be used to inform decisions on testing for COVID-19, clinical management of COVID-19, and workforce planning.

Our study has some limitations. First, reflecting the fact that the outbreak started from China, most studies were from China. However, given similar case-fatality rates and clinical manifestations across different countries, it seems likely that these results are largely generalizable. Nonetheless, we need to acknowledge regional variations of some risk factors (e.g., ∼25-fold difference in the prevalence of smoking in men vs. women in China^45^) and thus future investigations in different regions would be valuable. Second, we did not include non-English publications. Third, most studies reported odds ratios, which are known to overestimate risk ratio when the prevalence of exposures is relatively high. Fourth, we cannot deny the possibility that some patients were included in multiple studies especially in the China CDC report^17^ and other Chinese studies. Nonetheless, the pooled estimates were largely similar in analyses that excluded the China CDC data (data not shown). Finally, the literature of COVID-19 is growing rapidly, and thus there is a lag time from our literature search and publication.

On the other hand, our systematic review has several strengths: in-depth review of CVD and its risk factors, a clinically relevant definitions of severe COVID-19 that minimize subjective reporting, careful consideration of potential overlap of patients by conducting restrictive and inclusive meta-analyses, meta-regression to explore potential confounding, and relatively short elapsed time of ∼3 weeks between the literature search and manuscript submission.

In conclusion, our systematic review and meta-analysis found robust associations of older age and male sex as risk factors of severe COVID-19. Furthermore, despite the potential for confounding, our results suggest that hypertension, diabetes, and CVD are independently associated with severe COVID-19. These factors can be used to inform objective decisions on COVID-19 testing, clinical management, and workforce planning.

## Data Availability

This systematic review is based on published studies.

## Acknowledgements

This project is supported by Resolve to Save Lives, which is funded by Bloomberg Philanthropies, the Bill and Melinda Gates Foundation, and Gates Philanthropy.

